# Functional and structural biomarkers of cognitive outcomes after brain tumor resection

**DOI:** 10.1101/2020.09.01.20186205

**Authors:** Meritxell García, José Aguasvivas, Santiago Gil-Robles, Iñigo Pomposo, Lucia Amorouso, Manuel Carreiras, Ileana Quiñones

**Affiliations:** BCBL, Basque Center on Cognition, Brain and Language, San Sebastian, Spain; University of Basque Country (UPV/EHU), San Sebastian, Spain; BioCruces Research Institute, Bilbao Spain; Department of Neurosurgery, Hospital Universitario Quirónsalud, Madrid, Spain; IKERBASQUE. Basque Foundation for Science, Bilbao, Spain; University of the Basque Country, UPV/EHU, Bilbao, Spain

**Keywords:** glioma, resection, postoperative outcome, prediction, neuropsychological test, mapping

## Abstract

Cognitive explorations have demonstrated the activation of plastic mechanisms in slow-growing brain lesions, generating structural and functional changes. Due to its incidence, it is essential to investigate the reorganization of functional areas in brain tumor patients as well as formulating new approaches for predicting patient’s quality of life after tumor resection.

Following this perspective, we formulated an efficient methodology for postsurgical prognosis prediction, not only in terms of the structural damage but also to measure the neuroplastic changes associated with tumor appearence. Of note, most of previous studies employed a limited number of neuropsychological and clinical features for predicting patient prognosis. Our objective is to optimize the traditional model and to develop a method that can predict outcomes with high accuracy and identify the most significant features for cognitive impairment, working memory, executive control and language outcomes. Our approach is based on the inclusion of a large battery of neuropsychological tests as well as the introduction of grey and white matter morphological measures for model optimization. We employed Support Vector Machine (SVM), Decision Tree, and Naïve Bayes algorithms for testing the models and outcomes. Overall, SVM performance showed to be more accurate as compared to Decision Tree and Naïve Bayes. Specifically, we found that, by introducing connectivity variables (e.g., grey and white matter measures) Cognitive Status and Working Memory exhibited a predictive improvement. However, Language and Executive Control outcomes were not significantly predicted in none of the models. The importance of the present study resides in the employment of structural and functional variables for postsurgical outcome prediction. We found that connectivity variables are sensitive for predicting the postsurgical quality of life.

## Introduction

Brain tumors are abnormal growths of cells in the central nervous system. Depending on their nature, they may displace the brain parenchyma or infiltrate it, diffusing along the white matter tracts and invading healthy grey and white matter areas. Their incidence in Spain is 14.13/100,000, slightly over the median standard incidence in Europe since 2000 (taken from aecc). Brain tumors are typically detected after an epileptic seizure without any other evident cognitive deficit (Duffau, 2014). Intraoperative brain mapping during awake surgery is considered to be the *gold standard* technique for brain tumor resection, as it minimizes the risk for postoperative deficits. It improves the quality of life outcome of those patients post-surgically, while maximizing the tumor resection without damaging functional areas involved in the tumor lesion (Kim et al., 2009; Smith et al., 2008). Direct electrical stimulation (DES) is applied in awake surgeries while patients perform sensory-motor and language-related tasks to identify functional areas and avoid postoperative cognitive deficits (Duffau, 2017; Kong, Gibb, & Tate, 2016; Rofes et al., 2017). The use of this technique has led to a reduced mortality rate as a consequence of the tumor resection, but the quality of life of those patients is usually still affected (although to a lesser extent as compared to surgeries using general anesthesia).

*What will happen to me after the surgery? How I will feel after it? Will I have a normal life? Will I be able to go to work? How likely it is for me to have a good recovery? Will I have severe sequelae after the surgery?* These are some of the questions that worry and distress patients before undergoing brain surgery. They want to receive a clear prognosis about their quality of life after the surgery. However, there is still no efficient methodology able to provide an accurate estimation of their postsurgical outcome. Pursuing this purpose, it is crucial to develop tools that would allow experts to predict patients’ prognosis before surgery and that allow patients to deal with their anguish. The employment of a tool like this, based on inter-subject variability, would be useful not only for postsurgical prognosis, but also for surgical planning and for designing patient-tailored neurorehabilitation programs.

The critical period for postsurgical recovery, when the brain reorganizes itself to compensate for the lession, comprehends the first three months after the intervention (Duffau, 2005, 2006). By observing the plastic changes that take place during this period, our goal in the present study is to design a model that can predict postsurgical cognitive outcome and quality of life prognosis of each patient with the combination of demographic, behavioral and neuroimaging data. This model will be compared with more limited models, which are mainly focused on demographic and behavioral data.

To the best of our knowledge, there are only four articles proposing a model for postoperative outcome prediction (see Table 1). The most widely used variables are patient demographic variables (i.e. age, gender), tumor-related variables (i.e. type, location, laterality, volume, extent of resection) and clinical data (i.e. general cognitive tests, treatment, psychological state). Among the reviewed literature, the most significant features for cognitive outcome prediction are age, tumor location, tumor grade, tumor volume and extent of resection (Johnson, Sawyer, Meyers, O’Neill, & Wefel, 2012; Kim et al., 2009). Within these models, the most useful variables for predicting are the ones relative to the tumor characteristics and the age of the patient. Nonetheless, the studies that have employed a more extense battery including also neuropsychological tests have reported that attention, memory and executive function-related tests (e.g. Trail Making test, Verbal fluency, working memory) are the most predictive variables for postsurgical cognitive impairment (Klein et al., 2001; Talacchi, Santini, Savazzi, & Gerosa, 2011; A. S. Wu et al., 2011). Interestingly, all the available data has shown that general cognitive tests, such as Karnofsky Performance Score and MiniMental State Examination, are not sensitive for predicting postsurgical cognitive impairment (Johnson et al., 2012; Kaleita et al., 2004; Klein et al., 2001; Talacchi et al., 2011; Vergun et al., 2018; A. S. Wu et al., 2011).

**Table 1.**
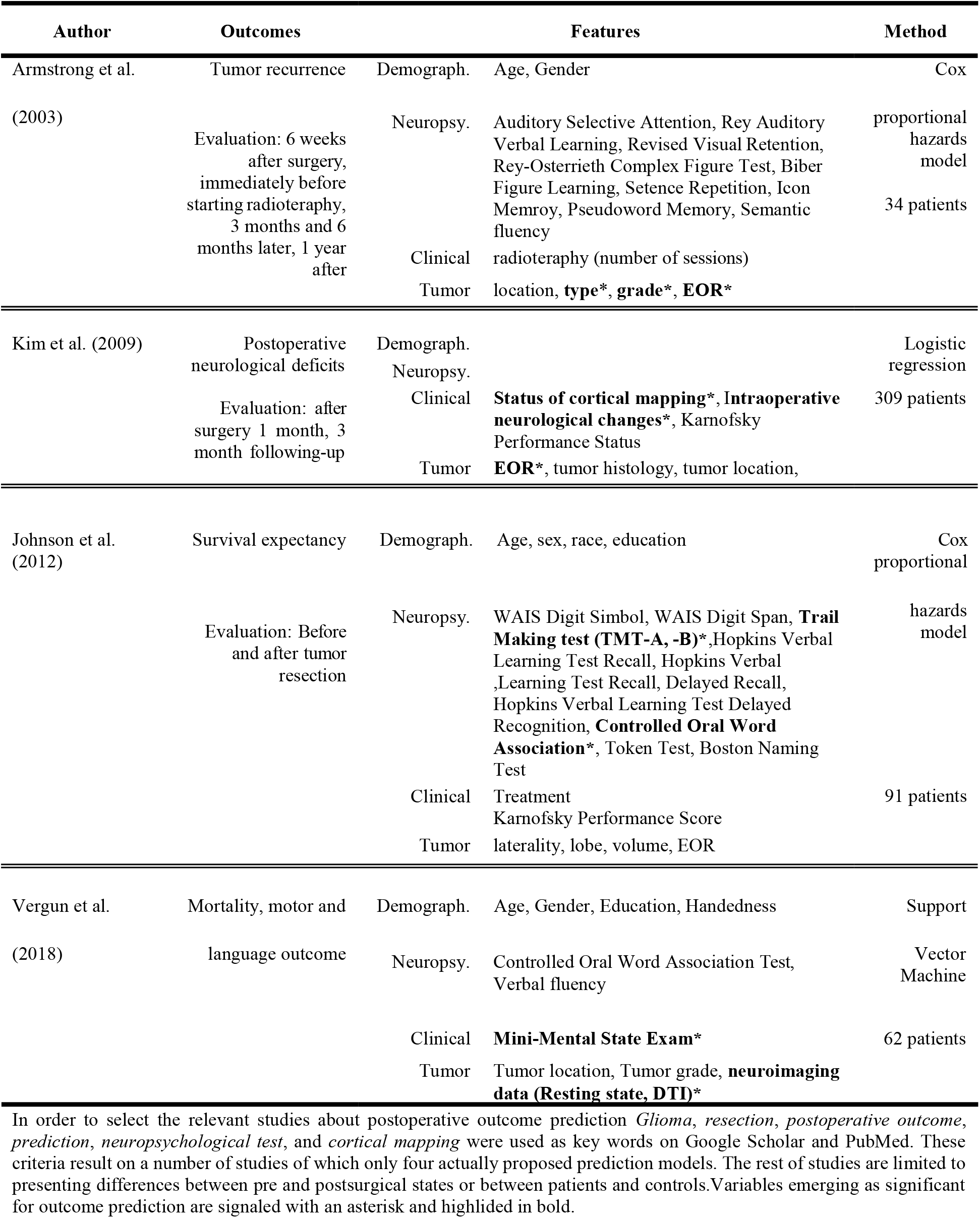
Summary of methodological approaches and results reported in the four previously published studies about postsurgical cognitive outcome prediction in patients with brain tumors.

Regarding the reviewed literature, the outcome prediction with this limited extent of tests may not be highly accurate. Our approach, conversely, relies on the implementation of an exhaustive battery of demographic variables, behavioral tests, tumor-related variables and clinical data, each one constituted by different features. In addition, whole-brain white and grey matter morphological measures have been introduced as crucial predictive features. All these neuropsychological tests and anatomical information are included as potential predictors.

In order to test the predictive capacity of each feature, we measured the accuracy of five models using three different machine learning algorithms (Support Vector Machine, Decision Tree and Naïve Bayes) and compared the level of significance independently. Secondly, in order to build upon the traditional model, we included more variables (language profile, extent of tumor volume in each specific region of grey and white matter, general and specific neuropsychological tests and subtests) as new features. Some of these neuroimaging features were included before (Vergun et al., 2018), demonstrating higher accuracy perfomance and prognostic capabilites than models based only on behavioral and demographic data. Our aim is to combine these two apporaches and to propose a model that detects significant features for predicting postsurgery cognitive deficits, language, working memory and executive control. This model will be supported by the employment of the three machine learning algorithms to predict patients outcome. For these reasons, we hypotesize that the inclusion of connectivity variables in the model, as whole brain grey and white matter measures, will increase the predicitive capacity of patients postsurgical outcome.

This developed methodology is expected to be of use in clinical practice, leading to direct implications towards neurorehabilitation not only in patients with brain tumors, but also in populations with similar acquired cognitive impairments (e.g., epileptic patients undergoing resection of their temporal lobe). Therefore, we developed this model to help surgeons from different domains, as it can be applied as a tool for surgery planning and for predicting patients’ outcome. In a further perspective, our objective is to expand this methodology to different clinical needs for cognitive deficit predictions and quality of life expectancy.

## Methods

### Participants

In the current study we drawed on a longitudinally test approach where a group of 13 patients (five females) with brain tumors were tested before and three months after the brain surgery. We acquired clinical, behavioral, and structural brain imaging data to extract potential predictors of postsurgery cognitive recovery. Patients with ages ranging from 22 to 42 years (mean = 40.0, standard deviation = 13.1) gave written informed consent to participate in this study as stipulated in the ethics approval procedure of the *BCBL Research Ethics Committee*. They all were native speakers of Spanish and had right-handed dominance, normal or corrected to normal vision and no history of psychiatric, or cognitive disabilities before the diagnosis. Patients were recruited at the Cruces Hospital in Bilbao, Spain, where they received their diagnosis and performed the surgery for tumour resection (see Table 2 for some individual features).

**Table 2.** Sociodemographic and Clinical Characteristics of Patients in the Study.

| Demographic characteristics |  |  |  |  | Clinical characteristics |  |  |
| --- | --- | --- | --- | --- | --- | --- | --- |
| No | Age | Gender | Education | LP | Tumor Type | Tumor Location | VL |
| 1 | 22 | 1 | 14 | 2 | Dysembryoplastic Tumour | Temp | 54 |
| 2 | 46 | 2 | 14 | 2 | Anaplastic astrocytoma Grade III | SMA | 0.55 |
| 3 | 44 | 1 | 14 | 1 | Glioblastoma Grade IV | Fus | 0.86 |
| 4 | 33 | 2 | 21 | 2 | Cavernoma | Fus | 2.54 |
| 5 | 54 | 2 | 17 | 1 | Cavernoma | SMA | 70.23 |
| 6 | 60 | 1 | 12 | 1 | Glioblastoma Grade IV | Fus | 5.8 |
| 7 | 31 | 2 | 20 | 1 | Grade II diffuse Astrocytoma | SMA | 16.45 |
| 8 | 13 | 1 | 6 | 1 | Anaplastic astrocytoma Grade III | IFG | 55.32 |
| 9 | 47 | 1 | 20 | 2 | High grade malignant tumor | SMA | 74.25 |
| 10 | 23 | 1 | 16 | 2 | Grade II Low Grade glioma | Temp | 1.85 |
| 11 | 56 | 1 | 12 | 2 | Oligodendroglioma Grade II | IFG | 53.92 |
| 12 | 48 | 2 | 17 | 1 | Glioblastoma Grade IV | SMA | 28.77 |
| 13 | 39 | 1 | 24 | 2 | Cavernoma | Temp | 96.52 |

LP = Language Profile; Temp = middle temporal gyrus extended into the anterior temporal pole; SMA = supplementary motor area extended into precentral regions and superior parietal areas; Fus = temporal areas affecting fusiform gyrus and medial temporal regions; IFG = inferior frontal gyrus extended into Insulas; VL = lesion volume (cm<sup>3</sup>).

### Neuro-psychological and behavioral assessment

Participants were assessed using a set of neuropsychological and behavioral tests providing us with a linguistic and a cognitive characterization of each individual. The screening consisted of different tests evaluating language abilities, verbal and non-verbal skills and overall cognitive status. Tests per cognitive domain are listed below:

- K-bit (Kaufman Brief Intelligence Test, Kaufman & Kaufman, 2013) test were employed as a measure of general intelligence.
- Working memory span, digit span, visuospatial memory, Cross-out, Stroop, and ANT were employed as measures of attention, working memory and executive function (Conway et al., 2005; Duncan-Johnson & Kopell, 1981; Redick & Engle, 2006).
- BEST (Basque, English, and Spanish Test) in L1 and L2 were employed as a measure of language proficiency (De Bruin, Carreiras, & Duñabeitia, 2017).
- Verbal and phonological fluency task (i.e., words retrieved by phonological or semantic cues) were used as a measure of fluency (Shao, Janse, Visser, & Meyer, 2014).
- PROLEC-SE-R (Cuetos, Arribas, & Ramos, 2016) were used as a measure of different language-specific operations (i.e., word and pseudoword reading; semantic associations, text comprehension, etc.).
- MMSE (Mini-Mental State Examination Folstein, Folstein, & McHugh, 1975) was used as a measured of cognitive impairment.

### MRI data: acquisition and analysis

Structural MRI for each participant were acquired at the BCBL using a Siemens 3T MAGNETOM Prisma Fit scanner. This system is equipped with a sixty four channel phased-array surface coil (Siemens, Erlangen, Germany), which provided a high spatial resolution and signal-to-noise ratio. T1-weighted MPRAGE anatomical volumes were acquired with the following parameters: echo time = 2.97 ms, repetition time = 2530 ms, flip angle = 7° and field of view = 256 × 256 × 160 mm3, number of axial slices = 176, slice thickness = 1 mm, in-plane resolution = 1 mm × 1 mm.

In order to obtained gray and white matter volumetric segmentations, T1 image data was pre-processed using the Voxel-Based Morphometry (VBM) toolbox (http://dbm.neuro.uni-jena.de/vbm.html) and the SPM12 software package. Images were corrected for bias-field inhomogeneity; tissue-classified into gray, white matter and cerebrospinal fluid; and registered to standard space using high-dimensional DARTEL normalization (the Diffeomorphic Anatomical Registration Through Exponentiated Lie Algebra, Ashburner, 2009). The segmentation approach used is based on an adaptive Maximum A Posterior technique, which does not need a priori information about tissue probabilities (Rajapakse et al., 1997). This procedure was further refined by accounting for partial volume effects and by applying a hidden Markov random field model which incorporates spatial prior information of the adjacent voxels into the segmentation estimation (Tohka et al., 2004). In order to measure regional differences in absolute volume of gray and white matter, the warped images were modulated. All the normalized-modulated images were smoothed with a filter of a 10 mm Gaussian kernel. In addition, a quality check was conducted for gray and white matter normalized-modulated images based on the sample homogeneity.

### Lesion analysis

3D lesion reconstruction were done by a trained technician using MRIcron software (Rorden, Karnath, & Bonilha, 2007) based on the combination of the T1-weighted MPRAGE, T2-weighted, and FLAIR images. A volume of interest was created for each patient each time point. Lesion overlap maps are displayed in Figures 1. From each pre and postsurgery 3D reconstruction, the tumor volume (cm^3^) was calculated. Additional measures of grey and white matter involvement were estimated. Extent of resection (cm^3^) was also measured on postoperative imaging as: Volume of (preoperative 3D Tumor Reconstruction ∩ postoperative Resection)*100 /preoperative tumor volume.

**Figure 1.**
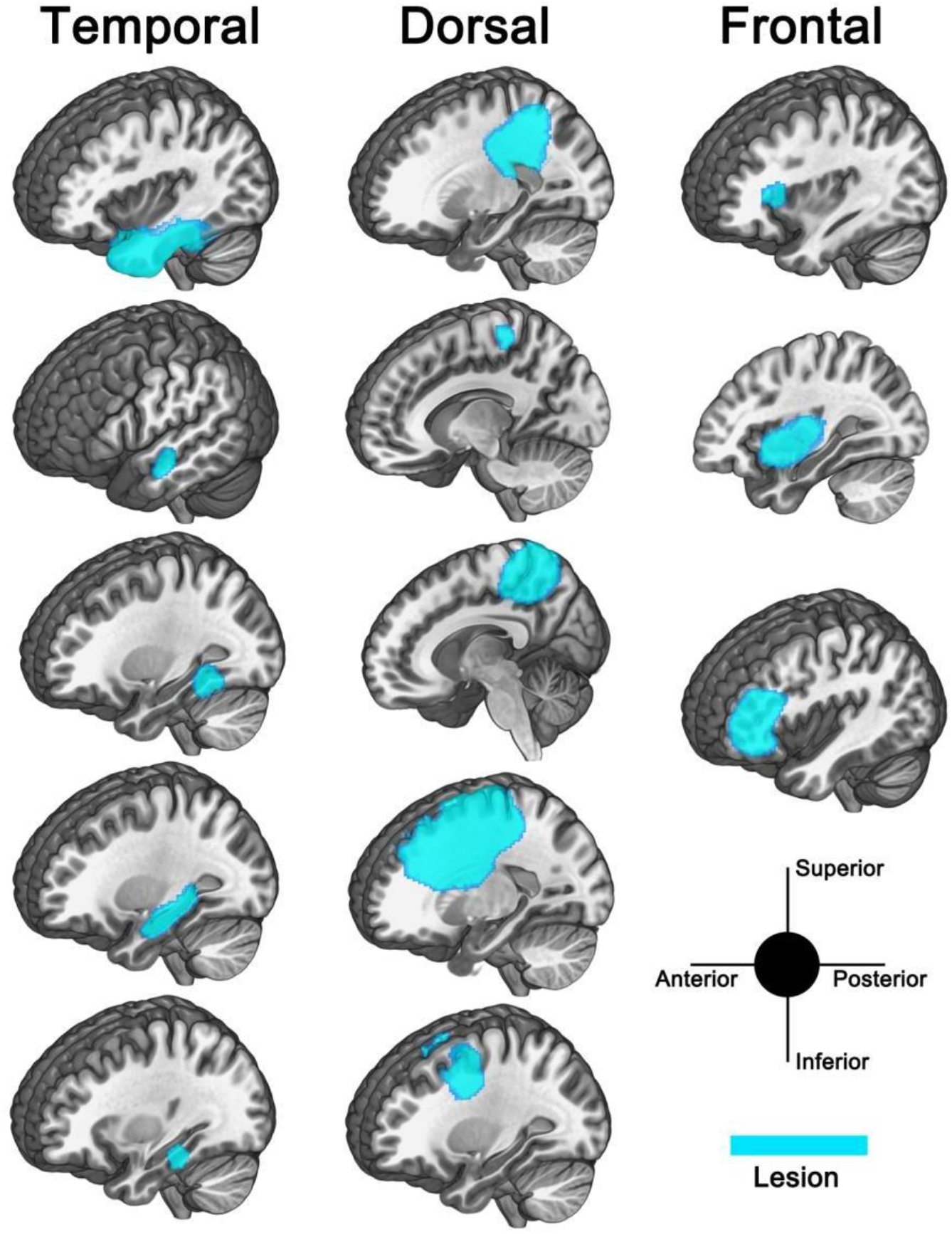
3D lesion reconstruction overlapped on the MNI single subject template.

### Data Preparation

The data comprised pre- and post-operative neuropsychological variables and demographic variables (i.e. age, gender, language profile, and education). The variables expected to be predicted by the model (outcomes) were excluded from the original list of features. Missing values were replaced with the median. In order to quantify the outcome of each variable, we created three categorical labels depending on the difference between the pre- and post-operative assessments. Namely, label 2 categorized the positive outcome, corresponding to performance improvement. The negative outcome, label 1, indicated a decrease in each task-related abilities and thus a decline in performance. Finally, label 0 indicated the preservation of the patient’s abilities. The outcome variables used as targets to predict were: *Working Memory*, obtained from the cross-out test’s scores; *Language* outcome, comprised of the vocabulary scores (K-bit); *Cognitive Status*, extracted from the MiniMental scores, and *Executive Control* composed of Stroop reaction times. None of the raw scores for these variables showed a significant correlation (all *p*s > 0.05).

### Machine Learning

Machine Learning (ML) is a popular technique that has been successfully applied in several domains nowadays. ML gives computers the ability to learn from data without explicitly being programmed for it (Samuel, 1988). Importantly, recent studies have employed ML for disease prediction and detection, suggesting that it could be an essential tool in clinical settings (Alanazi, Abdullah, & Qureshi, 2017; Claudino et al., 2019; Jiang et al., 2017). ML works with data itself rather than a given algorithm as traditional computing usually do. ML can be categorized to supervised and unsupervised learning. Supervised learning only needs input and output sample to return a model (algorithm) for predicting new data outcome. Unsupervised learning (Data Mining) needs only data input to find the correlation from the data input and to generate output and algorithm (Alonso-Betanzos & Bolón-Canedo, 2018). In this study, since we do not have sufficient data to arrive at the unsupervised learning requirement, we therefore focus on the Supervised ML methodologies.

Supervised ML algorithms are mainly used for regression and classification (Alanazi et al., 2017; Alonso-Betanzos & Bolón-Canedo, 2018). Regression is mostly used to solve continuous numerical problems by obtaining the most optimal fitting curve for the given data points that is able to predict new data accurately (i.e., house price or stock price prediction). Classification is made for treating discrete objective variables (i.e., categorical variables). It is widely used for forecast prediction, speller correction, search ranking of objectives, among other domains. Generally, supervised ML allows finding patterns in the input that maximizes the accuracy of outcome prediction (or minimizes the error between the predicted value and the target). This procedure is often accompanied by a pre-selection of the most relevant predictive features, which is the so-called feature selection. In order to avoid the over- or under-fitting problems, cross-validation techniques in which parts of the data are left as hold-out sets are crucial for evaluating and tuning the model.

In this study, we aim to build a classifier tool based on Supervised ML algorithms with our data. In the literature, many Supervised ML algorithms have been used for classification tasks (Claudino et al., 2019). We chose Support Vector Machine (SVMs), Decision Tree, and Naïve Bayes methods, which were reported as well-known algorithms for classification and prediction in Medicine and the Health Care domain (Alanazi et al., 2017; Jiang et al., 2017). For the most part, SVM algorithm has been successfully employed for classifying and predicting the recovery index of patients after brain surgery as well as evaluating pre-surgical language dominance (Gazit et al., 2016; Munsell et al., 2015). In this study, we compared SVM, Decision Tree, and Naïve Bayes capacity to predict post-surgical clinical outcomes

#### Comparison between different ML algorithms

SVM is a popular algorithm for binary classification and prediction (Alanazi et al., 2017; Platt, 1999; Smola & Schölkopf, 2004; T.-F. Wu, Lin, & Weng, 2004), owing to its high accuracy and low computational cost. Its popularity resides in the capacity for classifying different groups of inputs into two classes. The main idea of SVM is to find a hyperplane with the maximum margin distance between two groups of data points, and this binary classification can be also expanded to multi-class classification.

Unlike the SVM classification, Decision Tree does not require to find a hyperplane. Instead, it classifies the input into a space to build a tree in which every single class is represented by a leaf node. Its testing procedure is characterized by moving from the bottom of the tree, taking a single branch at each point. However, the computational complexity will depend on the dataset size (Breiman, 2017; Friedman, Hastie, & Tibshirani, 2001b).

Naïve Bayes is known as a probabilistic ML model used for classification objectives. It is based on the Bayes theorem and the assumption made here is that the features are independent, the presence of one particular feature has no effect on the other. It is performed using a maximum likelihood estimation method due to its parameter approximation mechanism (Alanazi et al., 2017; McCallum & Nigam). Contrarily to SVM, it can quickly reach a reasonable estimation of the parameters on small training datasets.

#### Outcome Prediction

All predictions were performed with the Scikit-learn package for Python3 (Vergun et al., 2018). All the features were scaled with respect to their maximum and minimum values. We tuned different classification algorithms to predict the outcome of clinical language categories. We have mainly focused on *Language*, *Working Memory*, *Executive Control*, and *Cognitive Status* as the predicted variables, due to their important role in representing the cognitive state of the patient both before and after the surgery (see section b).

#### Feature selection & tuning set

While a detailed characterization of each patient is clinically significant to identify fine-grained changes in both behavior and brain structure, one of the biggest problems in ML appears when the number of features is substantially larger than the number of samples (Vergun et al., 2018). To address this issue, the literature traditionally uses either theory-driven or data-driven approaches. Theory-driven approaches involve selecting a subset of the features based on previous work and specific hypotheses, while data-driven approaches encompass different methods for reducing the dimensionality based on the data at hand (e.g., Principal Component Analysis). One of the most widely used data-driven approaches is Recursive Feature Elimination (RFE), which reduces the dimensionality of the data based on each feature predictive ability. RFE fits a model and removes the weakest feature until a criterion is reached. In doing so, this method attempts to eliminate dependencies and collinearity that might bias the model’s prediction.

In this study, we employed RFE in combination with Leave-One-Out Cross-Validation (LOOCV) to tune the classifiers. LOOCV partitions the data into training and testing sets by holding out one sample as the test case, and performing the necessary tuning on the training data to try to predict this test case. This procedure allowed the selection of the most relevant features for the full 14 patient model. The procedure is analogous to that employed in Vergun et al. (Vergun et al., 2018).

#### Prediction and Accuracy

We employed LOOCV to estimate how accurate our predictive model was in practice. This method seems to be the best estimator of the models’ performance on an unseen patient and is preferred because it reduces under/overfitting issues (Friedman, Hastie, & Tibshirani, 2001a). Using this method, we obtained an accuracy score for each patient as a test case, which allowed us to calculate the overall accuracy and weighted average F1 scores of the five models for each outcome variable. The final accuracy was contrasted to chance level using one-sample t-test. F1 score indicates the weighted average between precision (positive predictive value) and recall (sensitivity), also taking into account the relative support (number of instances of that class) for each label (Sasaki, 2007). Overall, values closer to 1 indicate a better predictive performance of a model, while values closer to 0 suggest closer to chance-level prediction by the model.

#### Final datasets for prediction

The outcomes to be tested were Cognitive Status, Executive Control, Language, and Working Memory. We tested each of these outcomes by using five different models, each iteratively increasing the number of features in relation to the previous one. Model 1 (base model) is composed only of demographic and behavioral features (BehF), Model 2 includes behavioral features and tumor-related variables (BehF+TumorF), Model 3 includes the grey matter volume of the whole brain (BehF+TumorF+GMF). In the same scheme, Model 4 is similar to Model 3, but replacing the grey matter volume (GMF) by white matter fibre tracks affected in the entire brain (WMF). Finally, Model 5 involves all the features of the previous models (BehF+TumorF+GMF+WMF).

We used SVM, Decision Tree and Naïve Bayes algorithms to test their capacity to predict the outcomes mentioned above using each of the five models. This procedure resulted in a total of 20 feature-outcome tables per algorithm (a total of 60 training and testing sets). The idea was to test whether the increasing inclusion of features improved each algorithm’s ability to classify each outcome correctly.

## Results

Figure 1 shows the accuracy of each algorithm for each outcome and model. The following sections describe the results for the SVM algorithm, while section 6 provides an overview of the results for Decision Tree and Naïve Bayes. Additional information about the classification results can be found in Appendix 1, 2, and 3.

**Figure 1.**
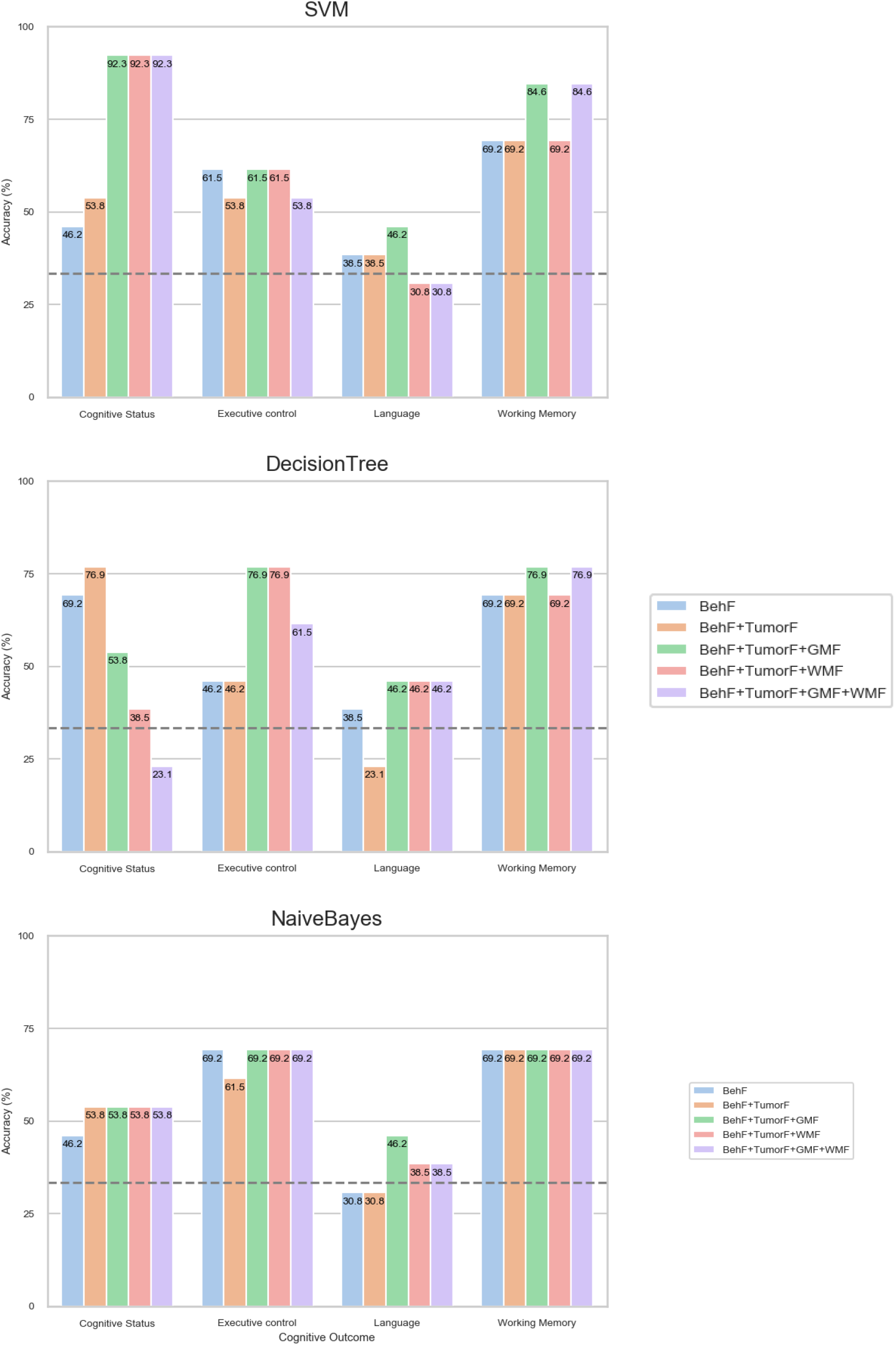
Prediction accuracy per algorithm, outcome, and model.

### Results for the SVM algorithm

#### Cognitive Status

Model 1 showed an accuracy of 46.15% (*p* = 0.39, F1 = 0.36). Although the accuracy improved to 53.85% by adding features from the Tumor in Model 2, it still did not reach significance (*p* = 0.18, F1 = 0.48). Models 3, 4, and 5 all showed significant accuracy of 92.31% (all *p*s < 0.01), suggesting that the addition of neuroimaging features contributed to the classification of cognitive status. The most common relevant features selected by the RFE process in Model 5 indicate a mix of feature levels, including behavioral (i.e., semantic fluency, gender, language profile), tumor (i.e., extent of resection), grey matter (i.e., left and right cerebellum, left cingulum, and left parahippocampal), and white matter (mainly the left and right superior longitudinal fasciculi).

#### Executive Control

Concerning Executive control, the models failed to reach a statistically significant result (all *p*s > 0.05). The best accuracy score of 61.54% was achieved by Models 1, 3, and 4.

#### Language

Results showed a similar trend for Language outcome, with none of the models achieving significant accuracy scores (all *p*s > 0.05). In this case, the highest accuracy was achieved by Model 3 (46.15%).

#### Working Memory

Model 1 showed a statistical significance accuracy of 69.23% (*p* = 0.02, F1 = 0.57) with only two relevant features (age and verbal fluency). Including the tumor features in Model 2 showed the same accuracy of 69.23% (*p* = 0.02, F1 = 0.57), even though some of the tumor features were selected as relevant features for classification (location and extent of resection). Models 3 and 5 achieved the highest accuracy of 84.62% (*p*s < 0.01, F1 = 0.81). Model 4 showed an accuracy of 69.23% (*p* = 0.02, F1 = 0.57). This suggests that the addition of white matter features did not improve the model above and beyond the behavioral and tumor features. Conversely, grey matter information, particularly in the cerebellum (left and right) and inferior parietal areas, appeared to increase the predictive accuracy of the SVM algorithm significantly.

### Results for the Decision Tree and Naïve Bayes algoritms

Decision Tree algorithm showed a seemingly inverse pattern for Cognitive Status compared to SVM, with accuracy decreasing as a function of the number of features. Model 2 achieved the highest accuracy of 76.92% (*p* < 0.01, F1 = 0.70). In contrast to SVM, Decision Tree showed significant accuracy in predicting Executive Control of 76.92% (*p* < 0.01, F1 = 0.77) with Models 3 and 4. However, none of the accuracies reached significance for Language outcome (all *p*s > 0.05). Finally, this algorithm showed a similar pattern in predicting Working Memory, with Models 3 and 5 showing the best predictive accuracy of 76.92% (*p* < 0.01, F1 = 0.76). Despite the somewhat comparable performance of Decision Tree and SVM, it is important to note that the number of features selected by Decision Tree were significantly larger than those selected by SVM.

In the case of Naïve Bayes, only Executive Control and Working Memory achieved significance, reaching an accuracy cap in Models 3, 4, and 5 of 69.23% (*p* = 0.02, F1 = 0.62). One possible explanation for such accuracy cap is rooted in Naïve Bayes difficulty to interpret collinearity in large feature sets due to its “naïve” approach. In this sense, it is possible that the algorithm is not suitable for RFE and subsequent classification. This is one of the reasons why Naïve Bayes appears to be the base classification algorithm in most papers (Alonso-Betanzos & Bolón-Canedo, 2018; Yang & Webb).

### Dissentangling language outcomes

As explained above, none of the employed algorithms managed to significantly predict language outcome as measured by vocabulary size scores in the K-bit test. Due to its importance for the quality of life, we also trained SVMs on other behavioral variables as measures of language outcome. We followed the same process as the previous outcomes of creating categories and excluding them as features from the datasets. Appendix 4 present the predictive capacity of SVM for predicting different language outcomes. Importantly, Model 5 showed an overall better predictive capacity for word reading and grammatical structure, while Models 3 and 4 showed their maximum accuracy for lexical selection. Model 4 also better predicted expositive comprehension, indicating an essential contribution of white matter features (mainly in inferior and superior longitudinal fasciculi).

## Discussion

The objective of this study was to develop a method that could be used for postsurgical prognosis, taking into account different types of outcomes that can affect quality of life. We expected that the addition of features with varying degrees of depth would lead to an increased predictive capacity for postsurgical quality of life. However, this hypothesis was only partially supported by the results, as they showed important contributions of adding grey and white matter features as predictors of Working Memory and Cognitive Status outcomes, but not for Language outcome and Executive Control.

This study represents a first step and a valuable contribution towards developing models that take into account distinct degrees of detail for pre-surgical patients. In our case, the degrees of detail ranged from behavioral measures to whole-brain accounts of structural grey and white matter. It extends previous literature regarding the prediction of post-surgical outcome by attempting to provide a rich characterization of each patient. In this regard, we first tested the predictive capacity of Models 1 and 2 in accordance to previous literature (Armstrong, Goldstein, Shera, Ledakis, & Tallent, 2003; Johnson et al., 2012; Kim et al., 2009; Talacchi et al., 2011). Both models, built from patient demographic information and clinical variables, are the ones widely tested in previous literature. These studies have traditionally proposed patients’ age and tumor location as the most significant variables for postsurgical prognosis (Armstrong et al., 2003; Habets et al., 2014; Klein et al., 2001). For instance, Kaleita et al. (Kaleita et al., 2004) postulated patients’ age and tumor location as the most significant indicators for neurocognitive functioning. Similarly, other papers presented other tumor related variables, such as tumor grade, volume, and extent of resection, as important factors in predicting postoperative cognitive state. In our study, however, these models only appeared to predict Working Memory outcome significantly, but not Cognitive Status. If we had limited the scope of this study only to models 1 and 2, we would have incorrectly concluded that post-operative cognitive decline can be evaluated and predicted (to some extent) by those variables.

Taking into consideration recent accounts on the plasticity ability of the whole brain in patients with brain damage, we introduced Models 3 and 4 to incorporate grey and white matter features, respectively. Both models seemed to accurately predict Working Memory and Cognitive Status above and beyond the initial models. Additionally, Model 5 introduced a combination of all features employed in the preceding models. The overall performance of this latter model for predicting Working Memory and Cognitive Status was as good as the ones with only white or grey matter. Furthermore, these models did manage to significantly predict specific language outcomes, such as grammatical structure, lexical selection, expositive comprehension, and word reading. Future studies could build upon these measures to develop sensitive composite scores to capture subtle variations in patient’s post-operative language outcome.

Previous studies have considered language impairments to be the ones most impacting on patients’ quality of life and have included them as the primary outcome to be predicted in the studies (Antonsson et al., 2018; Duffau, Gatignol, Mandonnet, Capelle, & Taillandier, 2008; Kim et al., 2009). Our results highlighted the importance of the inferior and superior longitudinal fasciculi, relevant for language production and comprehension, when introducing connectivity variables. These results are in line with (Duffau et al., 2008), where language connectivity and the overall grey and white matter organization were proposed as the main predictors of patients’ postsurgical cognitive outcomes. Despite the importance of the language organization in the brain, the present work is one of the only studies employing neuroimaging data as predicting features for quality of life prognosis.

To our knowledge, only (Vergun et al., 2018) used connectivity variables to predict post-surgical cognitive outcome. The fact that previous studies have not incorporated variables of connectivity in their research situates the present study as a novel approach for quality of life prognosis. At the same time, these findings elucidate the importance of considering whole-brain plasticity and not only contralateral areas as other studies have done (Armstrong et al., 2003; Habets et al., 2014). For these reasons, the present study establishes the need for optimizing predictive approaches by incorporating connectivity variables for a better quality of life prognosis. Moreover, the incorporation and optimization of the model proposed here will allow for a deeper understanding of cortical and subcortical network.

### Methodological importance

Our results showed that SVM generally performed better than the other two algorithms when applying them to our database. Given that SVM is assumed to give better results as the sample size increases (see Table 3), once we have a more extensive database, including more participants and more features, it should give a more accurate outcome prediction. In the long term, by increasing the database size, we would like to adapt the current “hot” methodology of deep learning algorithms (i.e., Neural Networks) into our models as well, and compare the performance between SVM and Neural Networks.

**Table 3.** Comparison of the three Machine Learning Algorithms employed: Support Vector Machine (SVM), Decision Tree, and Naïve Bayes.

| Algorithms |  |  |  |
| --- | --- | --- | --- |
|  | SVM | Decision Tree | Naïve Bayes |
| Advantages | Effective in high dimensional spaces | Computational complexity is not high | Still valid when dealing with small sample size |
|  | Suitable when the sample size is smaller than the number of dimensions | Output is easy to understand and to interpret (i.e., output tree can be visualized). | Easy to extend to multi-class classification problems |
|  | It offers various Kernel functions for non-linear decision boundaries | Can handle numerical and categorical data | Fast, efficient and easy to implement |
| Disadvantages | When the number of features larger than the number of samples, it is crucial to choose suitable Kernel function and regularization | Propensity to overfit the data | Sensitive to preprocessing of data input |
|  | Complex calculation when there are many class labels | Can be unstable because small variations in data might lead to different results | Only for categorized data |
|  | Not usually employed for continuous numerical variables, mostly for categorical variables | May generate biased tree if some classes are dominant |  |

During this project, our group has developed new scripts for implementing the machine learning algorithms and we have also developed Python script tools for general data collection purposes which can be useful for future work. For instance, the working memory script allows extracting data systematically from all the .log output files of the working memory task. Another example, the audio recognition script which benefits from Google speech recognition source, can interpret the audio files into words (English, Spanish and Basque are all available) and count the verb frequency automatically among hundreds of .wav files. All these scripts will be released in Github website as open-source tools.

### Study limitations

Despite the relevance of our results, some limitations need to be considered. One of the most determinant limitations is the small sample size. As it can be noticed, our final sample size only allows for an estimation of the models’ accuracy for the current dataset, thus limiting the generalizability to novel data. A direct implication of this factor is found in the statistical inference of each model. In some models, the three algorithms presented the same accuracy and standard deviation (given that there were only 13 samples to predict), which led to identical *p* values (see section 6). Motivated by the above reasons, future approaches must necessarily include a larger sample size.

Also, a point needs to be raised about the necessity to include more diversity in demographics and tumor histology variables. This would allow to increase the heterogeneity of our database and possibly lead to increasing generalizability of the results. Likewise, missing data is another barrier in the present study. In fact, a small sample size usually causes more difficulties when dealing with missing data, while with a larger database, this problem can be much more easily addressed.

## Conclusions

The current work has constructed a complete framework for predicting postsurgical quality of life in brain tumor patients. We have tested the predictive capacity of the traditional model and optimized it by introducing the effect of connective variables. Results show the importance of introducing grey and white matter fibre tracks information for predicting postoperative cognitive outcome. However, it is still not clear if grey and white matter are dependent or independent variables, due to the same statistical significance in the last two models. The present study is an approximation of a predictive methodology that has to be improved and further tested in larger samples to provide more conclusive results.

## Data Availability

Data included in this research have no availability

## Appendix 1

**Table 4.** Comparison between the different models obtained with SVM.

| Support Vector Machine (SVM) |  |  |  |  |  |  |  |
| --- | --- | --- | --- | --- | --- | --- | --- |
| Model | Data | Outcome | Accuracy | SD | *p* | F1 | No_features |
| Model 1 |  |  |  |  |  |  |  |
|  | BehF | Cognitive Status | 46.15 | 13.83 | 0.39 | 0.36 | 2 |
|  | BehF | Executive control | 61.54 | 13.49 | 0.07 | 0.47 | 2 |
|  | BehF | Language | 38.46 | 13.49 | 0.72 | 0.36 | 19 |
|  | BehF | Working Memory | 69.23 | 12.80 | **0.02** | 0.57 | 2 |
| Model 2 |  |  |  |  |  |  |  |
|  | BehF+TumorF | Cognitive Status | 53.85 | 13.83 | 0.18 | 0.48 | 6 |
|  | BehF+TumorF | Executive control | 53.85 | 13.83 | 0.18 | 0.43 | 2 |
|  | BehF+TumorF | Language | 38.46 | 13.49 | 0.72 | 0.36 | 29 |
|  | BehF+TumorF | Working Memory | 69.23 | 12.80 | **0.02** | 0.57 | 4 |
| Model 3 |  |  |  |  |  |  |  |
|  | BehF+TumorF+GMF | Cognitive Status | 92.31 | 7.39 | **<0.01** | 0.92 | 19 |
|  | BehF+TumorF+GMF | Executive control | 61.54 | 13.49 | 0.07 | 0.47 | 2 |
|  | BehF+TumorF+GMF | Language | 46.15 | 13.83 | 0.39 | 0.38 | 8 |
|  | BehF+TumorF+GMF | Working Memory | 84.62 | 10.01 | **<0.01** | 0.81 | 12 |
| Model 4 |  |  |  |  |  |  |  |
|  | BehF+TumorF+WMF | Cognitive Status | 92.31 | 7.39 | **<0.01** | 0.93 | 12 |
|  | BehF+TumorF+WMF | Executive control | 61.54 | 13.49 | 0.07 | 0.47 | 2 |
|  | BehF+TumorF+WMF | Language | 30.77 | 12.80 | 0.85 | 0.22 | 2 |
|  | BehF+TumorF+WMF | Working Memory | 69.23 | 12.80 | **0.02** | 0.57 | 2 |
| Model 5 |  |  |  |  |  |  |  |
|  | BehF+TumorF+GMF+WMF | Cognitive Status | 92.31 | 7.39 | **<0.01** | 0.92 | 16 |
|  | BehF+TumorF+GMF+WMF | Executive control | 53.85 | 13.83 | 0.18 | 0.43 | 3 |
|  | BehF+TumorF+GMF+WMF | Language | 30.77 | 12.80 | 0.85 | 0.22 | 2 |
|  | BehF+TumorF+GMF+WMF | Working Memory | 84.62 | 10.01 | **<0.01** | 0.81 | 9 |

F1= F score; No Features = number of features that are significant for each outcome.

## Appendix 2

**Table 4.** Comparison between the different models obtained with Decision Tree algorithm.

|  |  | Decision Tree |  |  |  |  |  |
| --- | --- | --- | --- | --- | --- | --- | --- |
| Model | Data | Outcome | Accuracy | SD | *p* | F1 | No_features |
| Model 1 |  |  |  |  |  |  |  |
|  | BehF | Cognitive Status | 69.23 | 12.80 | 0.02 | 0.64 | 8 |
|  | BehF | Executive control | 46.15 | 13.83 | 0.39 | 0.45 | 30 |
|  | BehF | Language | 38.46 | 13.49 | 0.72 | 0.37 | 11 |
|  | BehF | Working Memory | 69.23 | 12.80 | 0.02 | 0.69 | 30 |
| Model 2 |  |  |  |  |  |  |  |
|  | BehF+TumorF | Cognitive Status | 76.92 | 11.69 | <0.01 | 0.70 | 8 |
|  | BehF+TumorF | Executive control | 46.15 | 13.83 | 0.39 | 0.45 | 42 |
|  | BehF+TumorF | Language | 23.08 | 11.69 | 0.42 | 0.20 | 22 |
|  | BehF+TumorF | Working Memory | 69.23 | 12.80 | 0.02 | 0.69 | 2 |
| Model 3 |  |  |  |  |  |  |  |
|  | BehF+TumorF+GMF | Cognitive Status | 53.85 | 13.83 | 0.18 | 0.49 | 116 |
|  | BehF+TumorF+GMF | Executive control | 76.92 | 11.69 | <0.01 | 0.77 | 18 |
|  | BehF+TumorF+GMF | Language | 46.15 | 13.83 | 0.39 | 0.38 | 3 |
|  | BehF+TumorF+GMF | Working Memory | 76.92 | 11.69 | <0.01 | 0.77 | 2 |
| Model 4 |  |  |  |  |  |  |  |
|  | BehF+TumorF+WMF | Cognitive Status | 38.46 | 13.49 | 0.72 | 0.37 | 56 |
|  | BehF+TumorF+WMF | Executive control | 76.92 | 11.69 | <0.01 | 0.76 | 32 |
|  | BehF+TumorF+WMF | Language | 46.15 | 13.83 | 0.39 | 0.44 | 55 |
|  | BehF+TumorF+WMF | Working Memory | 69.23 | 12.80 | 0.02 | 0.72 | 109 |
| Model 5 |  |  |  |  |  |  |  |
|  | BehF+TumorF+GMF+WMF | Cognitive Status | 23.08 | 11.69 | 0.42 | 0.24 | 192 |
|  | BehF+TumorF+GMF+WMF | Executive control | 61.54 | 13.49 | 0.07 | 0.61 | 30 |
|  | BehF+TumorF+GMF+WMF | Language | 46.15 | 13.83 | 0.39 | 0.44 | 55 |
|  | BehF+TumorF+GMF+WMF | Working Memory | 76.92 | 11.69 | <0.01 | 0.77 | 175 |

F1= F score; No Features = number of features that are significant for each outcome.

## Appendix 3

**Table 6.**
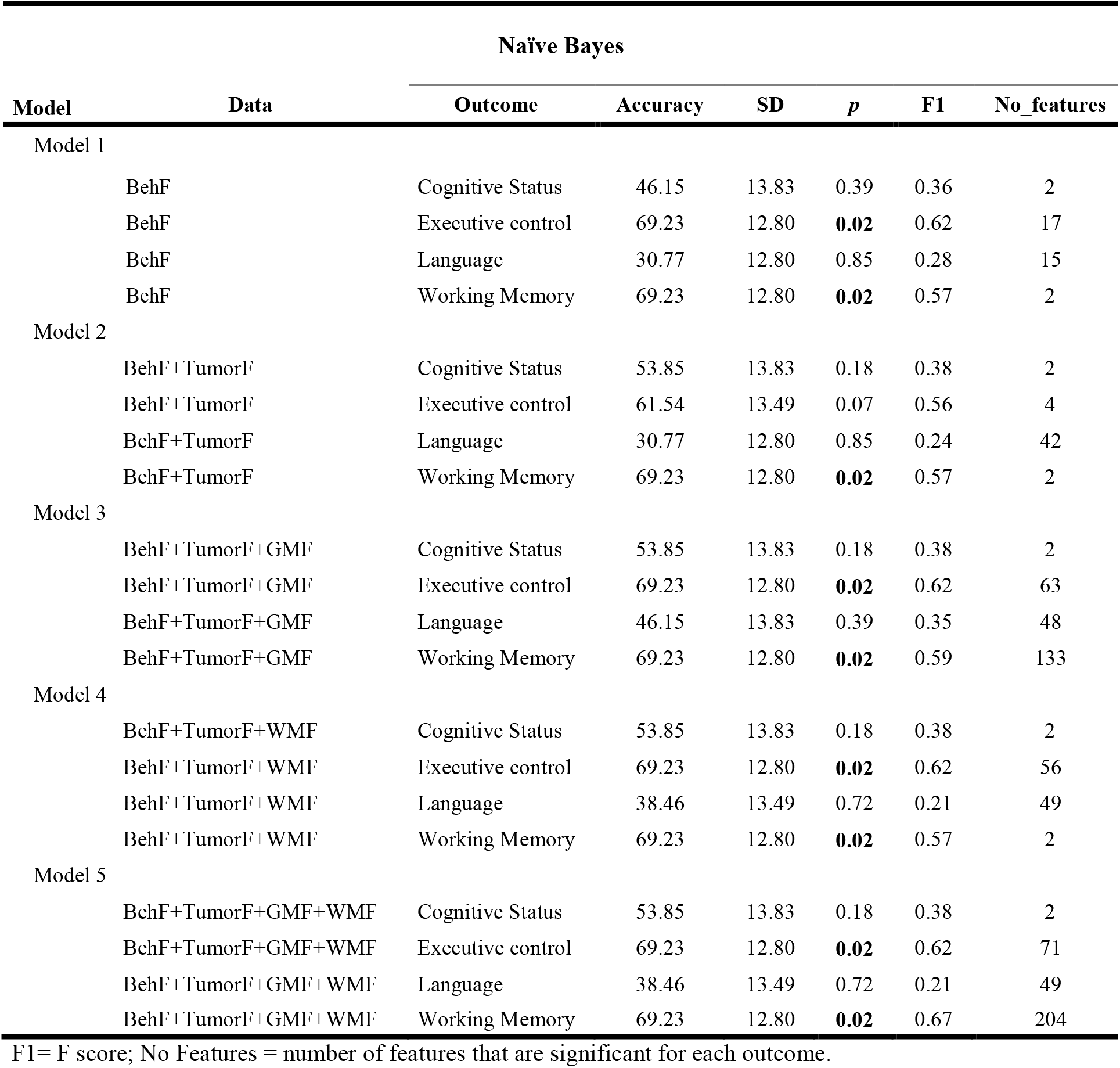
Comparison between the different models obtained with Naïve Bayes algorithm.

## Appendix 4

**Figure 2.**
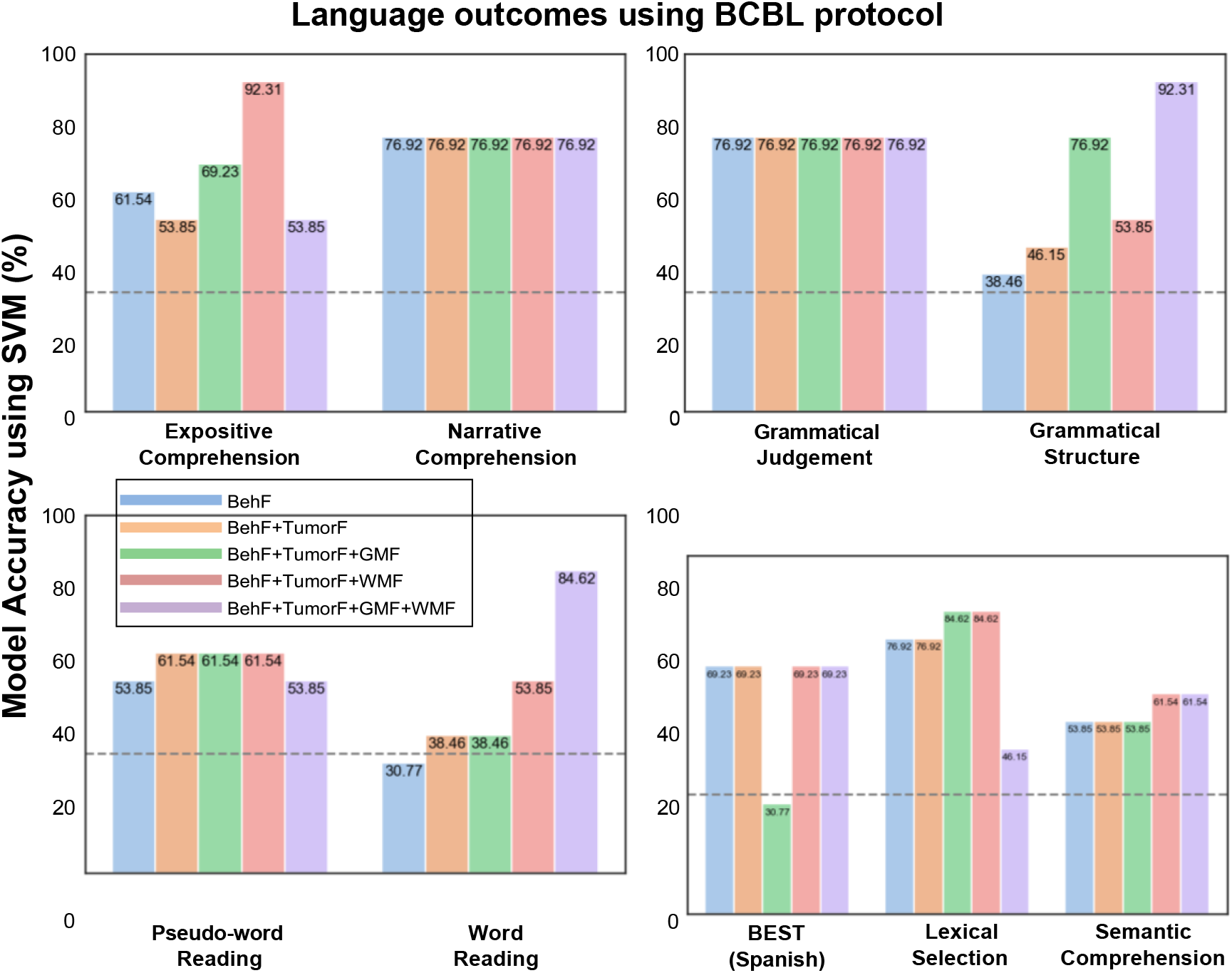
SVM prediciton accuracy in percentage for language outcome. Level of accuracy for each of the variables composing language outcome. 100% of accuracy means a reliability of that variable for language outcome.

